# Knowledge and practices towards COVID-19 among Palestinians during the COVID-19 outbreak: A second round cross-sectional survey

**DOI:** 10.1101/2020.10.13.20211888

**Authors:** Nouar Qutob, Faisal Awartani, Mohammad Asia, Imad Abu Khader

## Abstract

Coronavirus disease 2019 (COVID-19) is a highly transmissible illness that spreads rapidly through human-to-human transmission. To assess the knowledge and practices of Palestinians towards COVID-19 after the ease of movement restrictions, we collected data from Palestinian adults between June 15^th^ and June 30^th^ 2020. The participants’ pool represented a stratified sample of 1355 adults from Palestinian households across 11 governorates in the West Bank. The questionnaire included 7 demographic questions, 13 questions about participants’ knowledge and awareness of COVID–19, and 4 questions regarding the participants’ safety measures that had been taken in the last three months. Based on the results of this study, we conclude that the majority of participants have a good knowledge about COVID-19, but were not adequately committed to the infection control measures necessary to protect themselves and others. The findings may provide valuable feedback to lawmakers and health administrators to prevent the spread of the epidemic.

## Introduction

Coronavirus disease 2019, known as COVID-19, is an infectious respiratory disease caused by novel coronavirus SARS-CoV-2 [1] and is spread through human-to-human transmission by body fluid droplets from the mouth or nose, which can spread when a COVID-19 patients talk, coughs or sneezes. The virus can also spread through indirect contact with contaminated surfaces [2][3]. The common symptoms of COVID-19 include fever, dry coughing, and fatigue and may lead to more serious symptoms such as difficulty in breathing and chest pain [4].

Since its emergence in Wuhan, China in December 2019 [5], SARS-CoV-2 has spread rapidly around the globe and was declared by the World Health Organization (WHO) as a global pandemic [6], [7].

Daily, new cases and deaths are being reported worldwide[8]. Many countries have imposed lockdown and movement control. The effectiveness of these mitigation measures is highly dependent on cooperation and compliance of all members of society. Adherence to mandated protective measures is essential in stopping the spread of the virus but is also dependent on the population’s overall knowledge, attitudes and awareness, according to KAP theories[9]–[11]. Studies conducted during the SARS outbreak in 2003 suggest that knowledge, attitudes and practices towards viruses are associated with emotions among populations and can play an integral role in determining a society’s readiness to accept behavioral change measures from health authorities, especially that a gap still exists in the knowledge of the epidemiology, prevalence and clinical spectrum of Covis-19 [12]–[15].

A first round cross-sectional survey was conducted between April 19^th^ and May 1^st^ 2020, data was collected through Computer Assisted Phone Interviewing (CATI) using resident phone numbers available through the Reach Calling Center [16]. Here, we conducted a second round cross-sectional survey between June 15^th^ and June 30^th^ 2020, designed to assess the knowledge and practices towards COVID-19 among Palestinians after the ease of restrictions.

## Materials and Methods

### Participants

This cross-sectional study was conducted between June 15^th^ and June 30^th^ 2020.

The study involved 1355 participants from 11 governorates, including 112 localities (supplementary table 1). 1395 households were selected using 3-stage cluster sampling. The cluster of households or census track is considered to be a geographic location that is comprised of approximately 100 households. The process for conducting cluster sampling was carried as follows: (1) Selecting a cluster of households (2) Selecting 10 households randomly from each cluster and (3) Selecting a person at random from the selected household. The clusters were selected using probability proportional to size (PPS) sampling (Table 1).

**Table 1:**
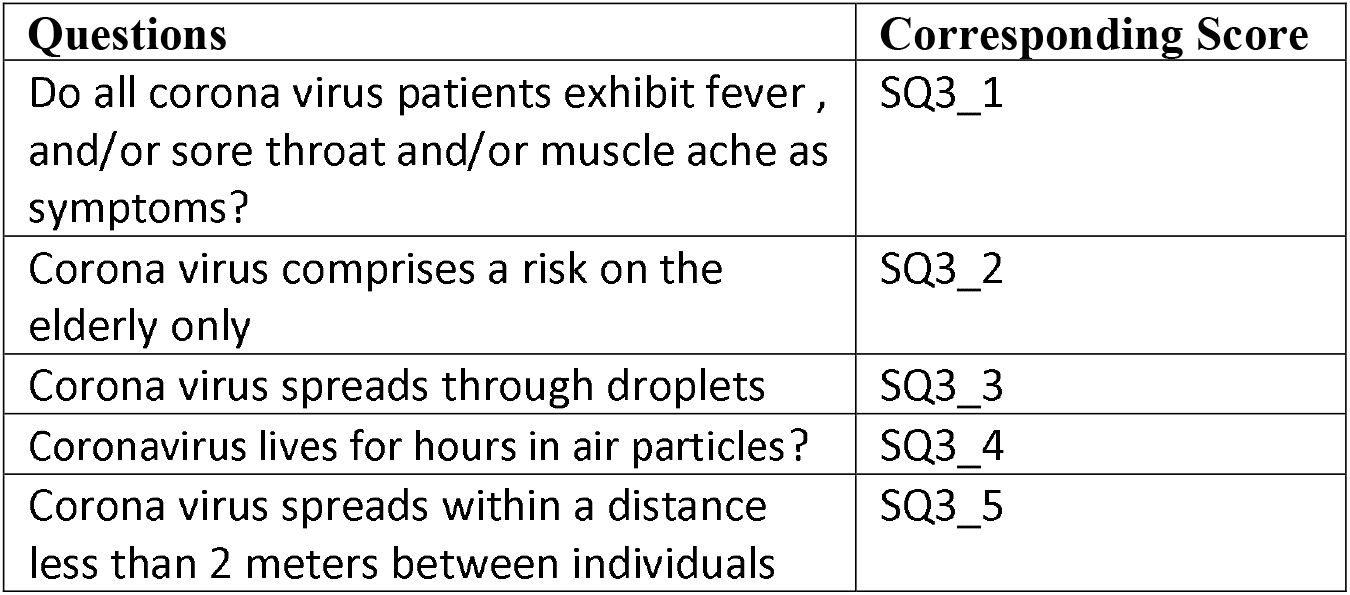
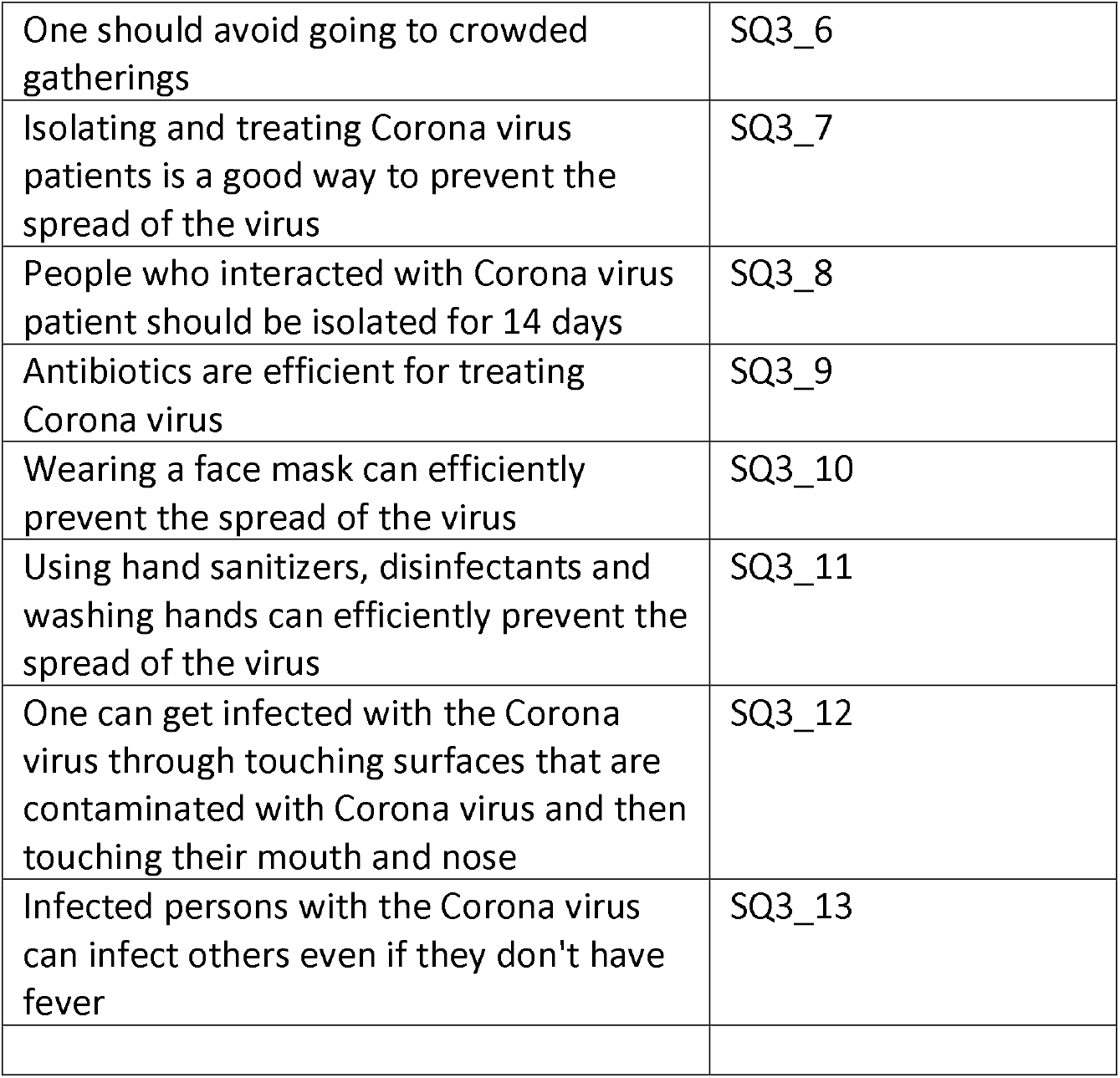
Questions used to test the participants knowledge about COVID-19

| Questions | Corresponding Score |
| --- | --- |
| Do all corona virus patients exhibit fever , and/or sore throat and/or muscle ache as symptoms? | SQ3_1 |
| Corona virus comprises a risk on the elderly only | SQ3_2 |
| Corona virus spreads through droplets | SQ3_3 |
| Coronavirus lives for hours in air particles? | SQ3_4 |
| Corona virus spreads within a distance less than 2 meters between individuals | SQ3_5 |
| One should avoid going to crowded gatherings | SQ3_6 |
| Isolating and treating Corona virus patients is a good way to prevent the spread of the virus | SQ3_7 |
| People who interacted with Corona virus patient should be isolated for 14 days | SQ3_8 |
| Antibiotics are efficient for treating Corona virus | SQ3_9 |
| Wearing a face mask can efficiently prevent the spread of the virus | SQ3_10 |
| Using hand sanitizers, disinfectants and washing hands can efficiently prevent the spread of the virus | SQ3_11 |
| One can get infected with the Corona virus through touching surfaces that are contaminated with Corona virus and then touching their mouth and nose | SQ3_12 |
| Infected persons with the Corona virus can infect others even if they don't have fever | SQ3_13 |

To select the number of clusters within each population location: (1) we calculated the sampling interval which equals the total number of households divided by the total number of clusters need to be selected by the sample say for example (m). The sampling interval SI= N/m, where N is the total number of households. (2) We selected a random number R0 between 0 and SI. (3) calculated Ri as R0+i*SI, a cluster is selected in Li if Ri belongs to the interval [Ci-1, Ci].

Field work was carried out between 15th^th^ June 2020 and 30^th^ June 2020 by a team of registered nurses, laboratory technicians, nursing students and laboratory technician students from the Arab American University following standardized health protocols (World Health, 2020).

Approval from National ethical committee was obtained (PHRC/HC/718/20). Written informed consents were obtained from the 1355 study participants.

## Measures

The questionnaire included 13 questions about participants’ knowledge and awareness of COVID–19 (Table 1). Since all questions were dichotomies (Yes/No) style, a correct answer was assigned 100 points and an incorrect answer was assigned 0 points. ANOVA test was used to calculate the significance.

The questionnaire also included 4 questions about the practices (Table 2). 100 points was assigned to full compliance and 0 points was given to lack of compliance within a given dimension.

**Table (2):**
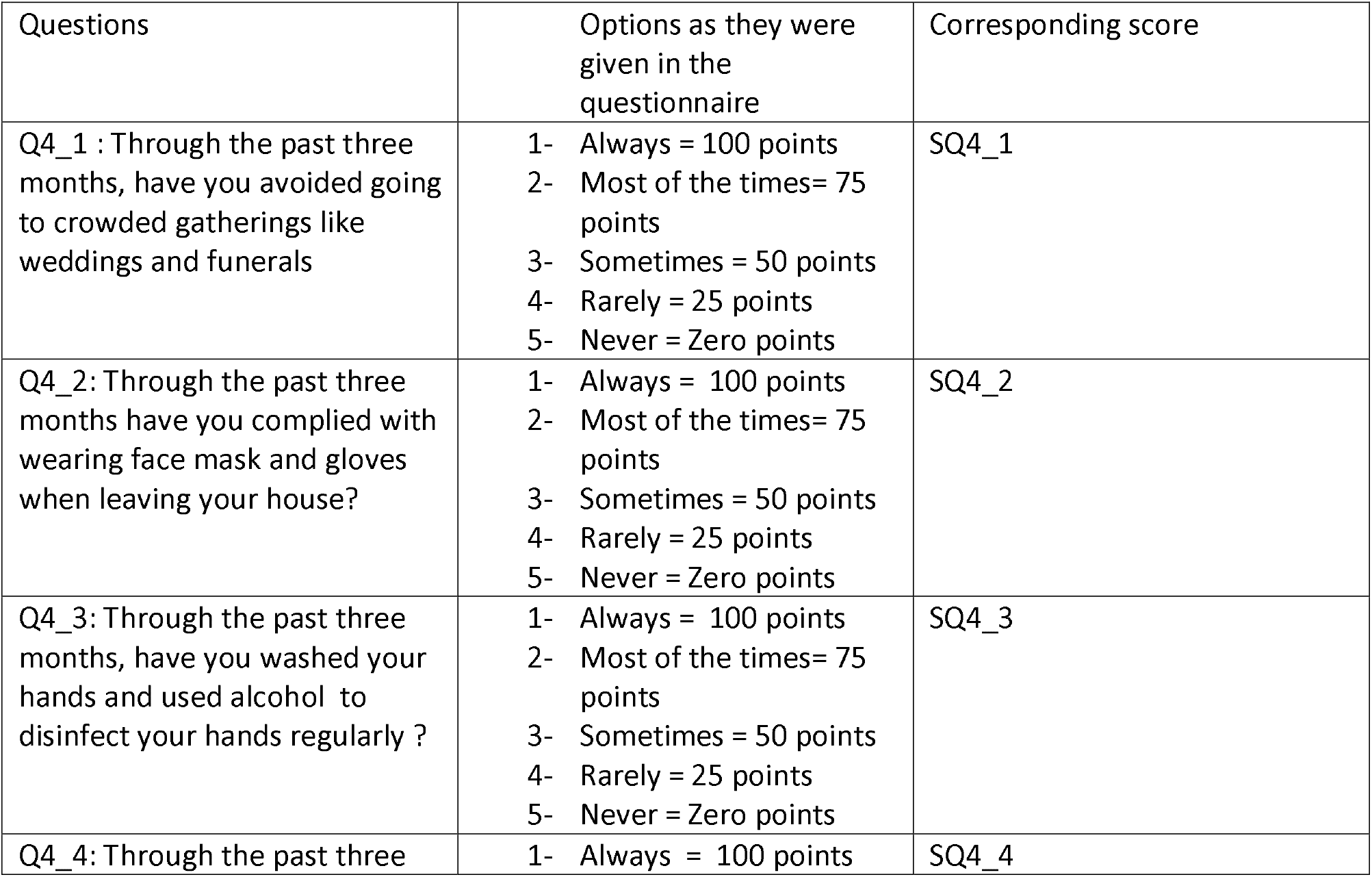
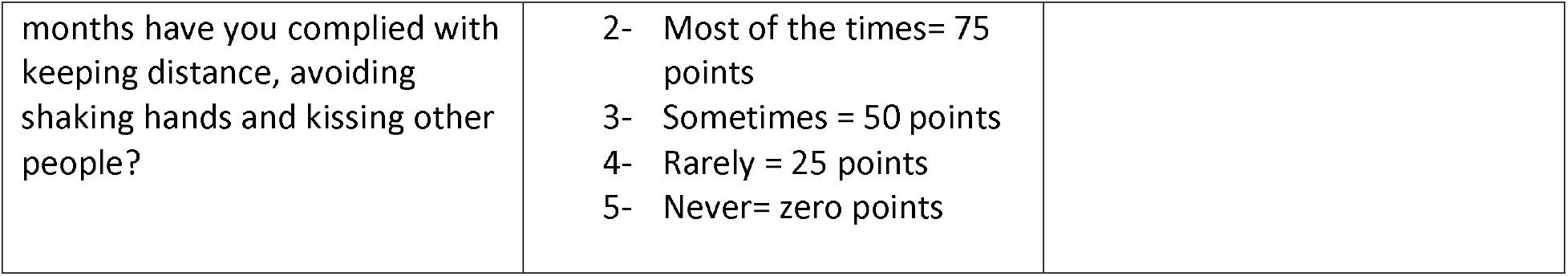
Questions used to measure the participants practices related to COVID-19 spread

| Questions | Options as they were given in the questionnaire | Corresponding score |
| --- | --- | --- |
| Q4_1 : Through the past three months, have you avoided going to crowded gatherings like weddings and funerals | 1- Always = 100 points<br>2- Most of the times= 75 points<br>3- Sometimes = 50 points<br>4- Rarely = 25 points<br>5- Never = Zero points | SQ4_1 |
| Q4_2: Through the past three months have you complied with wearing face mask and gloves when leaving your house? | 1- Always = 100 points<br>2- Most of the times= 75 points<br>3- Sometimes = 50 points<br>4- Rarely = 25 points<br>5- Never = Zero points | SQ4_2 |
| Q4_3: Through the past three months, have you washed your hands and used alcohol to disinfect your hands regularly ? | 1- Always = 100 points<br>2- Most of the times= 75 points<br>3- Sometimes = 50 points<br>4- Rarely = 25 points<br>5- Never = Zero points | SQ4_3 |
| Q4_4: Through the past three | 1- Always = 100 points | SQ4_4 |
| months have you complied with keeping distance, avoiding shaking hands and kissing other people? | 2- Most of the times= 75 points<br>3- Sometimes = 50 points<br>4- Rarely = 25 points<br>5- Never= zero points |  |

### Data Analysis

Data analysis was conducted using the statistical software (IBM SPSS 23.0) to produce a preliminary cross-tabulation of the study variables with the background indicators. This statistical report is comprised of cross-tabulations of all the study variables representing knowledge and practice as well as the demographic characteristics of respondents.

Knowledge Score was calculated as (SQ3_1+SQ3_2+…+SQ3_12+SQ3_13)/13 based on the dimensions given in table 1 and Practice score was calculated as (SQ4_1+SQ4_2+SQ4_3+SQ4_4)/4 based on the dimensions given in Table (2).

## Results

A total of 1355 participants from 1395 households across 11 governorates, including 112 localities (supplementary table 1). The sample included 26% in age group (15-24), 23% in age group (25-34), 28% in age group (35-49) and 23% tests in age group (50+). In total, the sample included 1218 males and 137 females.

We used ANOVA to test the bivariate relationship between knowledge score (dependent variable) and educational level as independent variable. The study showed an average knowledge score of 62 points out of 100 points. Our results show a statistically significant difference in knowledge between respondents who completed lower education vs those who completed higher education (P-value=0.048) (Table 3.A), and a statistically significant correlation between knowledge and age (p-value=0.008). The highest knowledge score (68 points) was reported among Palestinians with age category 50 – 59 years of age, while the lowest score (63 points) was among those with ages less than 39 years (Table 3.B). No significant difference was observed in females compared to males (P-value=0.53) (Table 3.C).

**Table 3.** A. 1. knowledge score of respondents who completed lower education versus those who completed higher education and 2. Anova results between and within groups. B. 1. knowledge score of respondents among different age groups and 2.Anova results between and within groups, C. 1 knowledge score of respondents among different genders and 2. Anova results between and within groups.

| Knowledge Scores | N | Mean | Std. Deviation | Std. Error | 95% Confidence Interval for Mean |
| --- | --- | --- | --- | --- | --- |

|  |  |  |  |  | Lower Bound | Upper Bound |
| --- | --- | --- | --- | --- | --- | --- |
| Preparatory or less | 374 | 61.6824 | 18.74460 | .96926 | 59.7765 | 63.5883 |
| Secondary | 483 | 61.4429 | 18.92001 | .86089 | 59.7513 | 63.1345 |
| College degree or more | 498 | 64.0408 | 16.50213 | .73948 | 62.5879 | 65.4937 |
| Total | 1355 | 62.4638 | 18.04538 | .49023 | 61.5021 | 63.4255 |

| Anova Results | Sum of Squares | df | Mean Square | F | Sig. |
| --- | --- | --- | --- | --- | --- |
| Between Groups<br>with Different<br>Education Levels | 1970.191 | 2 | 985.095 | 3.034 | .048 |
| Within Groups with<br>Different Education<br>levels | 438940.571 | 1352 | 324.660 |  |  |
| Total | 440910.762 | 1354 |  |  |  |

B
| Knowledge<br>Scores | N | Mean | Std. Deviation | Std. Error | 95% Confidence Interval for Mean |  |
| --- | --- | --- | --- | --- | --- | --- |
|  |  |  |  |  | Lower Bound | Upper Bound |
| 15-39 years | 505 | 61.4471 | 19.40679 | .86359 | 59.7504 | 63.1437 |
| 30- 39 years | 282 | 60.5565 | 19.19325 | 1.14294 | 58.3067 | 62.8063 |
| 40- 49 years | 253 | 64.2749 | 15.23110 | .95757 | 62.3890 | 66.1607 |
| 50- 59 years | 186 | 65.8395 | 15.67144 | 1.14909 | 63.5725 | 68.1065 |
| 60 + years | 129 | 62.1944 | 17.49448 | 1.54030 | 59.1466 | 65.2421 |
| Total | 1355 | 62.4638 | 18.04538 | .49023 | 61.5021 | 63.4255 |

| Anova Results | Sum of Squares | df | Mean Square | F | Sig. |
| --- | --- | --- | --- | --- | --- |
| Between Age<br>Groups | 4506.701 | 4 | 1126.675 | 3.485 | .008 |
| Within Age Groups | 436404.061 | 1350 | 323.262 |  |  |
| Total | 440910.762 | 1354 |  |  |  |

C.
| Knowledge Scores | N | Mean | Std. Deviation | Std. Error | 95% Confidence Interval for Mean |  |
| --- | --- | --- | --- | --- | --- | --- |
|  |  |  |  |  | Lower Bound | Upper Bound |
| Male | 1218 | 62.3595 | 18.16445 | .52047 | 61.3384 | 63.3806 |
| Female | 137 | 63.3914 | 16.98481 | 1.45111 | 60.5217 | 66.2610 |
| Total | 1355 | 62.4638 | 18.04538 | .49023 | 61.5021 | 63.4255 |

| Anova Results | Sum of Squares | df | Mean Square | F | Sig. |
| --- | --- | --- | --- | --- | --- |
| Between Gender Groups | 131.124 | 1 | 131.124 | .402 | .526 |
| Within Gender Groups | 440779.638 | 1353 | 325.779 |  |  |
| Total | 440910.762 | 1354 |  |  |  |

N: number of respondents; Std Deviation: *standard deviation*; Std. Error: standard error; df degrees of freedom; F: F-test.

The study indicates that the lowest knowledge score (10 points) was related to SQ3_1 which addresses knowledge of COVID-19 Symptoms, the second lowest score (29 points) was related to SQ3_2 addressing antibiotics as an efficient treatment for COVID-19 (table 4).

**Table 4:**
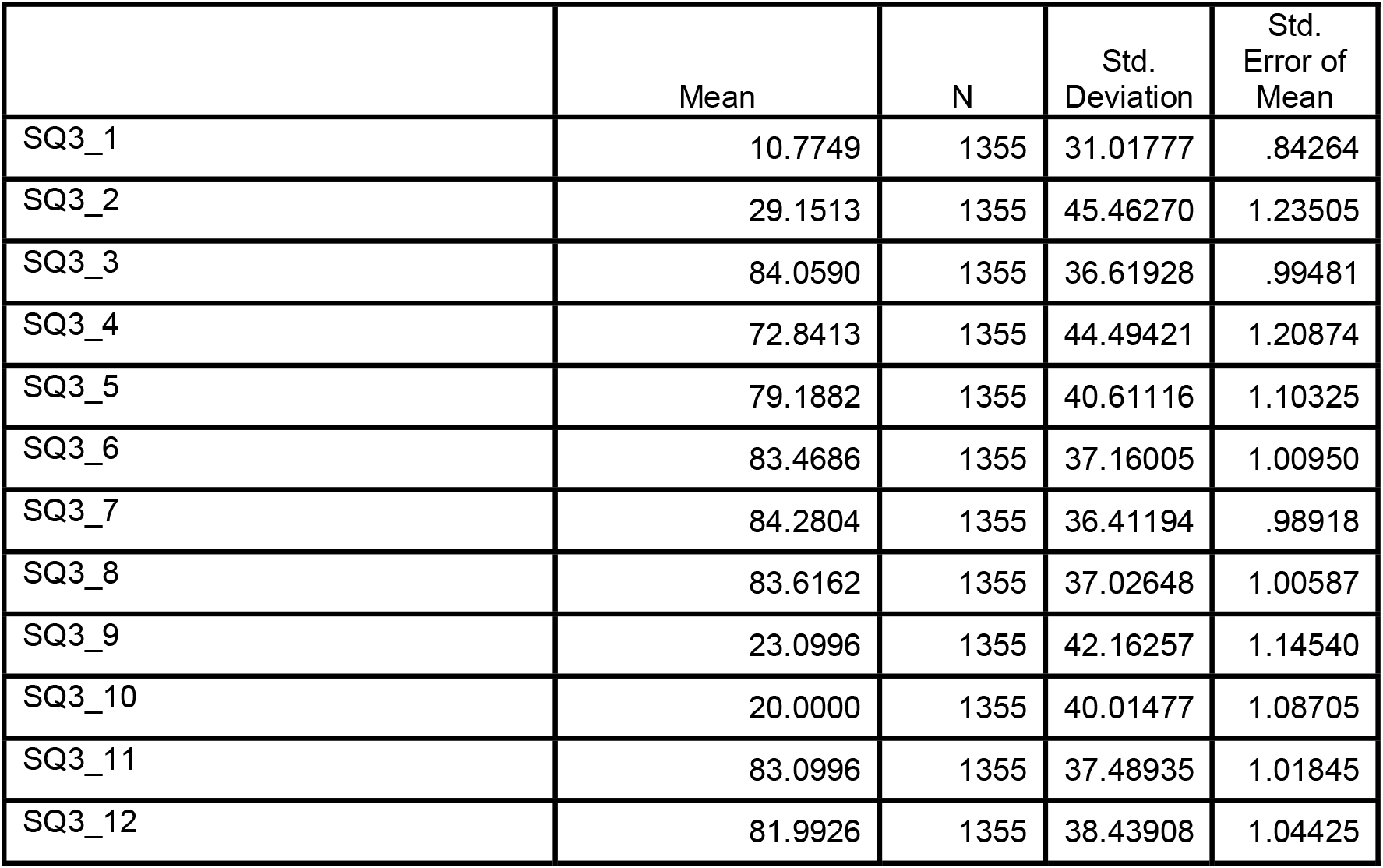
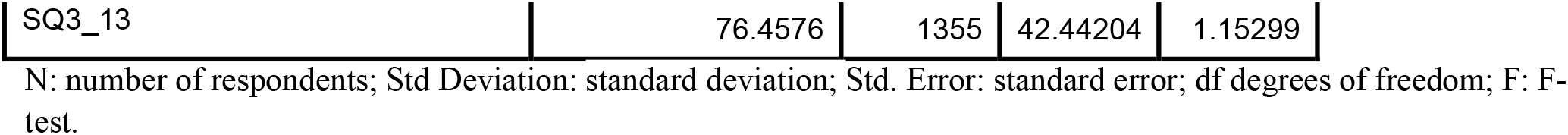
Mean score of knowledge variables

|  | Mean | N | Std. Deviation | Std. Error of Mean |
| --- | --- | --- | --- | --- |
| SQ3_1 | 10.7749 | 1355 | 31.01777 | .84264 |
| SQ3_2 | 29.1513 | 1355 | 45.46270 | 1.23505 |
| SQ3_3 | 84.0590 | 1355 | 36.61928 | .99481 |
| SQ3_4 | 72.8413 | 1355 | 44.49421 | 1.20874 |
| SQ3_5 | 79.1882 | 1355 | 40.61116 | 1.10325 |
| SQ3_6 | 83.4686 | 1355 | 37.16005 | 1.00950 |
| SQ3_7 | 84.2804 | 1355 | 36.41194 | .98918 |
| SQ3_8 | 83.6162 | 1355 | 37.02648 | 1.00587 |
| SQ3_9 | 23.0996 | 1355 | 42.16257 | 1.14540 |
| SQ3_10 | 20.0000 | 1355 | 40.01477 | 1.08705 |
| SQ3_11 | 83.0996 | 1355 | 37.48935 | 1.01845 |
| SQ3_12 | 81.9926 | 1355 | 38.43908 | 1.04425 |
| SQ3_13 | 76.4576 | 1355 | 42.44204 | 1.15299 |

N: number of respondents; Std Deviation: standard deviation; Std. Error: standard error; df degrees of freedom; F: F-test.

Our results showed a statistically significant difference in good practice between respondents who completed lower education vs those who completed higher education (P-value=0.000) (Table 5A), and a statistically significant correlation between practice and age (P-value=0.037). The lowest practice score (54 points) was among age group 15 – 29 years old, while the highest practice score (60 points) was among age group 30 – 39 years of age (Table 5B). (Table 3.B). Our data showed that practices score among Palestinians females (68 points) was higher than that among Palestinian males (59 points). The difference is statistically significant with P-value=0.004. (Table 5C).

**Table 5.** A. 1. Practice score of respondents who completed lower education versus those who completed higher education and 2.Anova results between and within groups with different educational levels. B. 1. Practice score of respondents among different age groups and 2.Anova results between and withinvage groups, C. 1.Practice score of respondents among different genders and 2.Anova results between and within gender groups.

| Knowledge Scores | N | Mean | Std. Deviation | Std. Error | 95% Confidence Interval for Mean |  |
| --- | --- | --- | --- | --- | --- | --- |
|  |  |  |  |  | Lower Bound | Upper Bound |
| Preparatory or less | 374 | 56.2166 | 34.80002 | 1.79947 | 52.6782 | 59.7549 |
| Secondary | 483 | 57.2593 | 33.22763 | 1.51191 | 54.2886 | 60.2301 |
| College degree or more | 498 | 65.7882 | 28.58226 | 1.28080 | 63.2717 | 68.3046 |
| Total | 1355 | 60.1061 | 32.33360 | .87838 | 58.3829 | 61.8292 |

| Anova Results | Sum of Squares | df | Mean Square | F | Sig. |
| --- | --- | --- | --- | --- | --- |
| Between Groups with Different Education Levels | 25650.622 | 2 | 12825.311 | 12.476 | .000 |
| Within Groups with Different Education levels | 1389904.440 | 1352 | 1028.036 |  |  |
| Total | 1415555.062 | 1354 |  |  |  |

**B.**
| Knowledge Scores | N | Mean | Std. Deviation | Std. Error | 95% Confidence Interval for Mean |  |
| --- | --- | --- | --- | --- | --- | --- |
|  |  |  |  |  | Lower Bound | Upper Bound |
| 15-29 years | 505 | 56.8317 | 33.33639 | 1.48345 | 53.9172 | 59.7462 |
| 30- 39 years | 282 | 63.4309 | 31.11793 | 1.85305 | 59.7832 | 67.0785 |
| 40- 49 years | 253 | 62.5988 | 31.69802 | 1.99284 | 58.6741 | 66.5236 |
| 50- 59 years | 186 | 59.2742 | 31.87037 | 2.33685 | 54.6639 | 63.8845 |
| 60 + years | 129 | 61.9671 | 32.06011 | 2.82274 | 56.3818 | 67.5523 |
| Total | 1355 | 60.1061 | 32.33360 | .87838 | 58.3829 | 61.8292 |

| Anova Results | Sum of Squares | df | Mean Square | F | Sig. |
| --- | --- | --- | --- | --- | --- |
| Between Age Groups | 10679.249 | 4 | 2669.812 | 2.566 | .037 |
| Within Age Groups | 1404875.813 | 1350 | 1040.649 |  |  |
| Total | 1415555.062 | 1354 |  |  |  |

**C.**
| Knowledge Scores | N | Mean | Std. Deviation | Std. Error | 95% Confidence Interval for Mean |  |
| --- | --- | --- | --- | --- | --- | --- |
|  |  |  |  |  | Lower Bound | Upper Bound |
| Male | 1218 | 59.2570 | 32.34675 | .92684 | 57.4386 | 61.0754 |
| Female | 137 | 67.6551 | 31.33167 | 2.67685 | 62.3615 | 72.9487 |
| Total | 1355 | 60.1061 | 32.33360 | .87838 | 58.3829 | 61.8292 |

| Anova Results | Sum of Squares | df | Mean Square | F | Sig. |
| --- | --- | --- | --- | --- | --- |
| Between Gender Groups | 8685.480 | 1 | 8685.480 | 8.353 | .004 |
| Within Gender Groups | 1406869.582 | 1353 | 1039.815 |  |  |
| Total | 1415555.062 | 1354 |  |  |  |

N: number of respondents; Std Deviation: *standard deviation*; Std. Error: standard error; df degrees of freedom; F: F-test.

Our study shows that the first two practices dimensions SQ4_1 which addresses the avoidance of crowded places and SQ4_2 which addresses compliance with wearing face masks got the lowest scores (54 points and 57 points respectively). SQ4_3 which addresses compliance with washing hands on a regular basis and SQ4_4 which addresses compliance with social distancing got higher scores (Figure 1).

**Figure 1:**
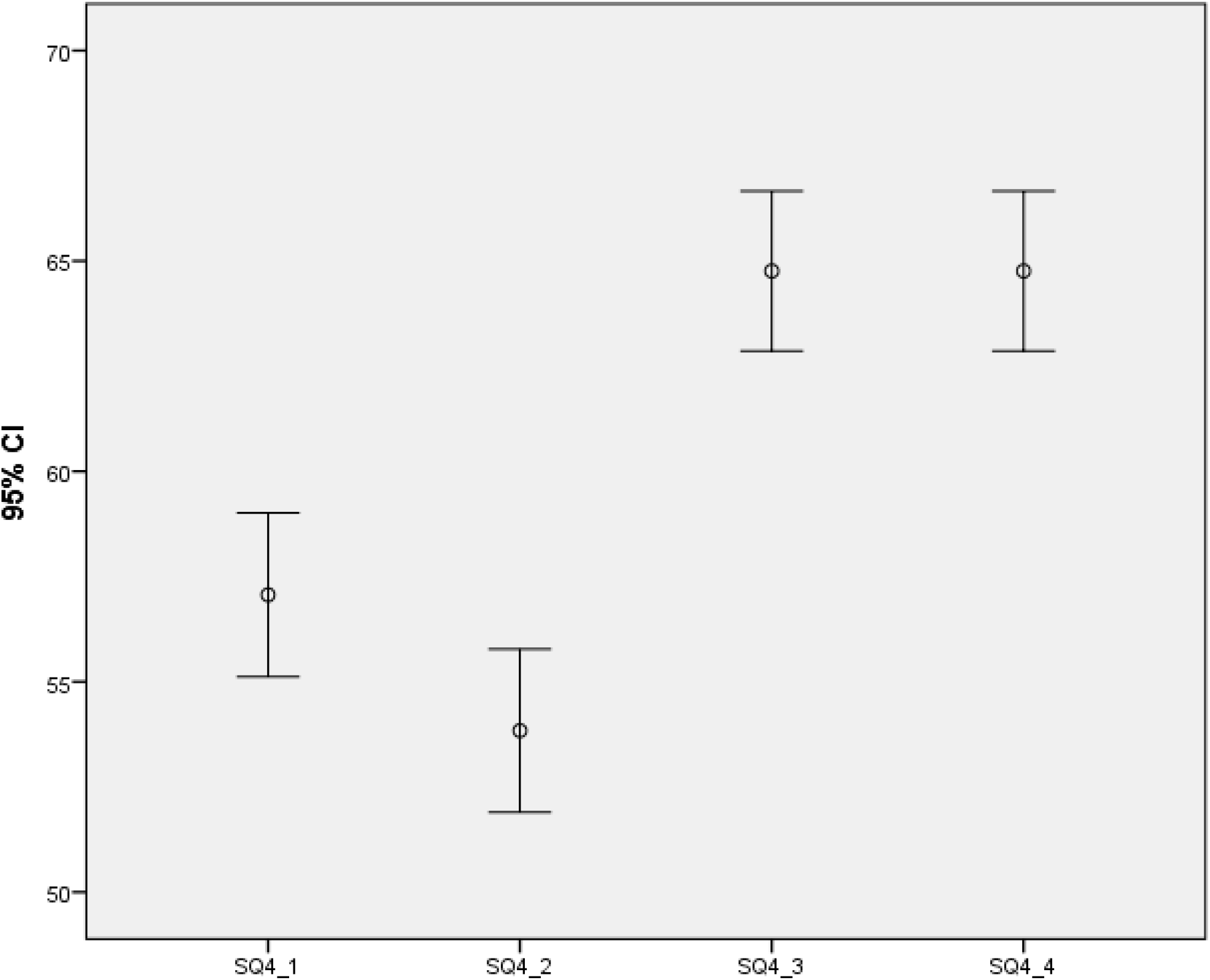
Mean score of the four dimensions comprising the practice score

## Discussion

This is a second round cross-sectional survey done in Palestine to examine the knowledge and practices towards COVID-19 among Palestinians.

In our study, we found that the majority of participants had a good base of knowledge about COVID-19, which is consistent with other studies conducted worldwide [14], [17] and with our first round study [16]. However, there was a low practice of health measurements among the participants.

Our results indicate that participants with higher educational level are more knowledgeable and apply better practices than those with lower educational level. This is consistent with a study conducted in Jordan, Saudi Arabia and Kuwait which reported high education level as an important predictor of greater COVID-19 knowledge scores (p<0.01) [18].

The differences between the two scores were statistically significant with P-value=0.048, 0.000 respectively. On another note, Our study also shows a statistically significant correlation between knowledge and age as well as good practice and age (p-value=0.008, p-value=0.037, respectively). The lowest knowledge score was among those with ages less than 39 years and practice score was among age group 15 – 29 years old. This result implies that efforts should be put to draw target groups and implement COVID-19 education on people with lower educational level and age groups less than 39 years old. These findings clearly indicate the importance of drawing target groups and implementing COVID-19 knowledge of targets via health education and media as this may both improve their outlook, motivate individuals to make appropriate decisions and result in safer personal practices, especially that good practice of hand hygiene, regular use of masks and gloves, social and physical distancing and self-quarantine have proven to limit prevalence of many infectious diseases and reducing the widespread infections [19], [20].

Our data shows that even though there is no significant difference between females and males in terms of their knowledge about COVID-19, practices score among Palestinians females is higher than that among Palestinian (P-value=0.004). This may be a consequence of females experiencing higher levels of stress, anxiety, and depression [21]. A survey conducted by KFF found that women are more likely than men to worry about the negative consequences of the pandemic and to report mental health effects from worrying about coronavirus. The study reported that women stay at home instead of going to work, school, or other regular activities [22]. In this respect, women tend to maintain social distancing and play an essential role in public health management [23].

Lastly, our study shows that the first two practices dimensions which address the avoidance of crowded places compliance with wearing face masks got low knowledge scores, indicating the need to educate a promote Palestinians to adhere to social distancing measures in order to stop the spread of COVID-19.

In conclusion, our study provides valuable feedback to lawmakers working to stop the spread of the virus. Conducting cross sectional surveys on a regular basis is important to facilitate the implementation of effective policy by enabling health officials to better understand the knowledge and practices of the Palestinian population towards COVID-19.

## Data Availability

All data will be available upon request

## Acknowledgments

We thank the participants for their cooperation. We thank the registered nurses, laboratory technicians, nursing students and laboratory technician students from the Arab American University: Ahmad Hodrub, Mohammad Faisal, Wajdi Tu ‘ma, Adam Maraw’a, Sharhabeel Nasrallah, Hisham Zahran and Mohammad Barakat.

## Funding Resources

The study was supported by the Arab American University.

